# N95 Mask Decontamination using Standard Hospital Sterilization Technologies

**DOI:** 10.1101/2020.04.05.20049346

**Authors:** Anand Kumar, Samantha B. Kasloff, Anders Leung, Todd Cutts, James E. Strong, Kevin Hills, Gloria Vazquez-Grande, Barret Rush, Sylvain Lother, Ryan Zarychanski, Jay Krishnan

**Affiliations:** Sections of Critical Care Medicine and Infectious Diseases, Departments of Medicine, Medical Microbiology and Pharmacology, University of Manitoba, Winnipeg Canada; National Microbiology Laboratory, Public Health Agency of Canada, Winnipeg Canada; National Centre for Foreign Animal Diseases, Canadian Food Inspection Agency, Winnipeg Canada; Section of Critical Care Medicine, Department of Medicine, University of Manitoba, Winnipeg Canada; Sections of Critical Care and Hematology, Departments of Medicine and Community Health Sciences, University of Manitoba, Winnipeg Canada

## Abstract

The response to the COVID-19 epidemic is generating severe shortages of personal protective equipment around the world. In particular, the supply of N95 respirator masks has become severely depleted, with supplies having to be rationed and health care workers having to use masks for prolonged periods in many countries. We sought to test the ability of 5 different decontamination methods: autoclave treatment, ethylene oxide gassing, low temperature hydrogen peroxide gas plasma treatment, vaporous hydrogen peroxide exposure and peracetic acid dry fogging to decontaminate a variety of different N95 masks of experimental contamination with SARS-CoV-2 or *Vesicular stomatitis virus* as a surrogate. In addition, we sought to determine whether masks would tolerate repeated cycles of decontamination while maintaining structural and functional integrity. We found that one cycle of treatment with all modalities was effective in decontamination and was associated with no structural or functional deterioration. Vaporous hydrogen peroxide, peracetic acid dry fogging and autoclave treatments were associated with no loss of structural or functional integrity to a minimum of 10 cycles for the mask models tested. The molded N95 masks however tolerated only 1 cycle of autoclaving. The successful application of autoclaving for layered fabric, pleated masks may be of particular use to institutions globally due to the virtually universal accessibility of autoclaves in health care settings.

---

The COVID-19 pandemic is proving to be an exceptional stress on hospital and health systems resources around the world. Many countries are experiencing or imminently expecting shortages for a variety of equipment and disposable supplies. A tightening supply of N95 masks that allow for protection from airborne pathogens and aerosolized viruses including SARS-CoV-2 is of particular and immediate concern. Without an adequate supply of N95 masks, health care providers are at extreme risk of contracting COVID-19. The occurrence of patient to health care worker spread of SARS-CoV-2 at sufficiently high rates would lead to demoralization of the workforce, depletion of health care workers for quarantine and would turn hospitals into hotspots for infection transmission.

N95 masks are normally single use products. However, according to news reports, re-use of N95 masks is ongoing in multiple institutions in the United States, Italy, Spain and India. Persistent shortages may increase the re-use of N95 masks globally as the pandemic progresses.

We sought to determine whether a range of different N95 masks would retain structural and functional integrity after treatment with widely available standard hospital decontamination techniques. Concurrently, we also determined the ability of each decontamination technique to effectively inactivate virus on experimentally inoculated masks.

## Methods

Several different N95 respirator masks were assessed using standard autoclaving, vaporous hydrogen peroxide (VHP) exposure, peracetic acid dry fogging system (PAF), ethylene oxide (EtO) gassing and low temperature hydrogen peroxide gas plasma (LT-HPGP) treatment.

The masks utilized were 3M’s VFlex 1804, Aura 1870, 1860, 8210 and 9210 respirator models (3M Company, St. Paul, Minnesota) as well as AO Safety 1054S (Pleats Plus) Respirator (Aearo Company, Indianapolis).

Standard autoclaving was performed using an Amsco Lab 250 model (Steris Life Sciences, Mentor, OH) with a peak temperature of 121 C for 15 min; total cycle time was 40 min (10 min conditioning/air removal, 15 min exposure, 15 min drying/exhaust).

EtO gas treatment was done using the model 5XLP Steri-Vac Sterilizer/Aerator (3M Company, St. Paul, Minnesota) with 1 hr exposure and 12 hr aeration time.

Low temperature hydrogen peroxide gas plasma (LT-HPGP) treatment was performed using a STERRAD^®^ 100NX sterilizer (Advanced Sterilization Products, Irvine, California). This device generates hydrogen peroxide vapor from 59% liquid H_2_O_2_, which is then electromagnetically excited to a low-temperature plasma state. Highly reactive species are generated from the hydrogen peroxide vapor in this state to facilitate faster decontamination of medical equipment. A standard 47-minute cycle was used for the mask treatment.

Vaporous hydrogen peroxide (VHP) treatment was performed with the VHP^®^ ARD System (Steris, Mentor, OH), it uses 35% liquid H_2_O_2_ to generate hydrogen peroxide vapor. A one hour program cycle was used, which consisted of 10 min dehumidification, 3 min conditioning (5 g/min), 30 min decontamination (2.2 g/min) and 20 min aeration. Peak VHP concentration was 750 ppm.

For peracetic acid fogging, a dry fogging system using fogger head and nozzles purchased from Ikeuchi USA (Blue Ash, OH) was used as described elsewhere [1]. One tenth diluted Minncare Cold Sterilant, a liquid peracetic acid (Mar Cor Purification, Skippack, PA) was used. The fogger was run until the relative humidity rose to 80-90%, which required 30 ml of the diluted chemical. The fogger was then turned off and the masks exposed for 1 hr.

VHP and dry fogging fumigation treatments were conducted in a 40 ft^3^ glovebox (Plas Labs Inc. Lansing, MI).

### Effectiveness of decontamination

The ability of each decontamination technology to inactivate infectious virus was assessed using experimentally inoculated masks. Small swaths cut from one of each of the 4 respirator models was surface contaminated on the exterior with *Vesicular stomatitis virus*, Indiana serotype (VSV) or SARS-CoV-2 (contaminated group). SARS-CoV-2 was only utilized if the decontamination method was done within the CL3 suite at Canada’s National Microbiology Laboratory. VSV was used if the decontamination method was only available at our hospital. The inoculum was prepared by mixing the virus in a tripartite soil load (bovine serum albumin, tryptone, and mucin) to mimic body fluids. 10 µl of the resulting viral suspension containing 6.75 log TCID_50_ (VSV) or 6.5 log TCID_50_ (SARS-CoV-2) was spotted onto the outer surface of each respirator at 3 different positions. Following 1-2 hr of drying, masks were placed into each of the decontamination devices. Similarly prepared and dried mask swaths were processed for virus titer determination to account for the effect of drying on virus recovery.

Following decontamination, virus was eluted from the mask material by excising the spotted areas on each mask swath and transferring each into 1 ml of virus culture medium (DMEM with 2% fetal bovine serum and 1% penicillin-streptomycin). After 10 minutes of soaking and repeated washing of the excised material, the elution media was serially diluted in virus culture medium for evaluation in a fifty-percent tissue culture infective dose (TCID_50_) assay. 100 µl of each dilution was transferred into triplicate wells of Vero E6 cells (ATCC CRL-1586) seeded 96 well plate. At 48 hours post-infection, cells were examined for determination of viral titres via observation of cytopathic effect. Titres were expressed as TCID_50_/ml as per the method of Reed and Muench [2].

### Impact of decontamination on structural and functional integrity

A group of the N95 masks without viral contamination (clean group) underwent multiple decontamination treatments by all the decontamination methods. Afterwards, these respirator masks were visually and tactilely assessed for structural integrity and underwent quantitative fit testing using a TSI PortaCount 8038+ to assess functional integrity. Masks were considered to be functionally intact if quantitative fit testing resulted in a fit factor of more than 100 for normal and deep breathing exercises [3]. For autoclaving, VHP, and PAF we assessed integrity after 1, 3, 5 and 10 cycles; for LT-HPGP treatment after 1, 2, 5 and 10 cycles; and for EtO gas treatment after 1 and 3 cycles.

## Results

### Effectiveness of Decontamination

All the decontamination treatments validated here have successfully inactivated the challenge VSV from any of the four mask materials in comparison to the untreated drying controls (Table 1). As a result, a demonstrable reduction of greater than six logs of infectious virus was recorded for all treated masks.

**Table 1:** Sterilization Efficacy of Decontamination Methods.

| **Mean virus recovery post-decontamination (LogTCID<sub>50</sub> ± SD) compared to drying controls** |  |  |  |  |  |  |  |
| --- | --- | --- | --- | --- | --- | --- | --- |
| Virus | Mask | Control | Autoclave | VHP | PAF | EtO | LT-HPGP |
| VSV | 3M 1860 | 6.1 ± 0.3 | 0 | 0 | 0 | 0 | 0 |
|  | 3M Aura 1870 | 6.5 ± 0.8 | 0 | 0 | 0 | 0 | 0 |
|  | 3M Vflex 1804 | 6.4 ± 0.2 | 0 | 0 | 0 | 0 | 0 |
|  | AO Safety 1054 | 6.5 ± 0.3 | 0 | 0 | 0 | 0 | 0 |
| SARS-CoV-2 | 3M 1860 | 5.2 ± 0.6 | 0 | 0 | 0 | ND | ND |
|  | 3M Aura 1870 | 5.2 ± 0.6 | 0 | 0 | 0 | ND | ND |
|  | 3M Vflex 1804 | 6.0 ± 0.7 | 0 | 0 | 0 | ND | ND |
|  | AO Safety 1054 | 6.3 ± 0.7 | 0 | 0 | 0 | ND | ND |

ND = not done, 0 = no growth
PAF: peracetic acid dry fogging system, VHP = vaporous hydrogen peroxide
LT-HPGP = low temperature hydrogen peroxide gas plasma
Control: small swaths of masks were inoculated with virus on their exterior and allowed to dry for 1-2 hr

Mask materials inoculated with SARS-CoV-2 had no recoverable virus following autoclaving, VHP and peracetic acid dry fogging treatments (Table 2). The titer of the starting SARS-CoV-2 virus was slightly lower than that of VSV, therefore the demonstrated log reduction ranged from 5.2-6.3. We could not validate the effectiveness of EtO and LT-HPGP against SARS-CoV-2 as they were not available at the National Microbiology Laboratory.

**Table 2:** Quantitative Fit Testing Results Of N95 Masks After Repeat Decontamination Cycles.

| PortaCount Result (normal & deep breathing exercises only) |  |  |  |  |  |  |
| --- | --- | --- | --- | --- | --- | --- |
| Groups |  | Masks |  |  |  |  |
| Control |  | 3M 1860 | pass |  |  |  |
|  |  | 3M Aura 1870 | pass |  |  |  |
|  |  | 3M Vflex 1804S | pass |  |  |  |
|  |  | AO Safety 1054S | pass |  |  |  |
|  |  | 3M 8210 | pass |  |  |  |
|  |  | 3M 9210 | pass |  |  |  |
|  |  |  | # of cycles |  |  |  |
|  |  |  | 1 | 3 | 5 | 10 |
| Autoclave |  | 3M 1860 | pass | fail | fail | fail |
|  |  | 3M Aura 1870 | pass | pass | pass | pass |
|  |  | 3M Vflex 1804S | pass | pass | pass | pass |
|  |  | AO Safety 1054S | pass | pass | pass | pass |
|  |  | 3M 8210 | pass | fail | fail | fail |
|  |  | 3M 9210 | pass | pass | pass | pass |
|  |  |  | 1 | 3 |  |  |
| EtO |  | 3M 1860 | pass | pass |  |  |
|  |  | 3M Aura 1870 | pass | pass |  |  |
|  |  | 3M Vflex 1804S | pass | pass |  |  |
|  |  | AO Safety 1054S | pass | pass |  |  |
|  |  |  | 1 | 2 | 5 | 10 |
| LT-HPGP |  | 3M 1860 | pass | fail | fail | fail |
|  |  | 3M Aura 1870 | pass | fail | fail | fail |
|  |  | 3M Vflex 1804S | pass | fail | fail | fail |
|  |  | AO Safety 1054S | pass | pass | fail | fail |
|  |  | 3M 8210 | pass | fail | fail |  |
|  |  | 3M 9210 | pass | fail | fail |  |
|  |  |  | 1 | 3 | 5 | 10 |
| VHP |  | 3M 1860 | pass | pass | pass | pass |
|  |  | 3M Aura 1870 | pass | pass | pass | pass |
|  |  | 3M Vflex 1804S | pass | pass | pass | pass |
|  |  | AO Safety 1054S | pass | pass | pass | pass |
|  |  |  | 1 | 3 | 5 | 10 |
| PAF |  | 3M 1860 | pass | pass | pass | pass |
|  |  | 3M Aura 1870 | pass | pass | pass | pass |
|  |  | 3M Vflex 1804S | pass | pass | pass | pass |
|  |  | AO Safety 1054S | pass | pass | pass | pass |

In summary, all decontamination methods resulted in no growth of virus in decontaminated specimens.

### Impact of decontamination on structural and functional integrity

All decontamination methods resulted in preserved structural and functional integrity of masks for at least one cycle of treatment (Table 2). The 3M Vflex 1804 models exhibited some mild bleeding of the ink label upon autoclaving but there was no impact on function. Autoclaving resulted in functional failure of the 3M 1860 and 8210 (molded) models after the first cycle but the other masks (all layered fabric, pleated), retained integrity through 10 cycles, the highest number tested. All masks treated with EtO retained integrity though 3 cycles (maximum number of cycles tested). LT-HPGT-treated masks failed testing beyond the first cycle, while VHP and PAF treatments maintained mask integrity through the maximum 10 cycles tested.

## Discussion

The unprecedented nature of the COVID-19 epidemic has revealed previously unrecognized deficiencies in global pandemic preparedness. In particular, the depletion of single-use disposable personal protective equipment has resulted in considerable health care worker anxiety and prolonged use of gear far beyond standard recommendations. The international shortage of N95 masks, which protect against exposure to aerosolized virus (which may occur during intubation and other invasive tracheobronchial procedures), is of particular concern given the respiratory nature of the SARS-CoV-2 infections. The shortage of these masks and their use for periods beyond recommended may be part of the reason for the reported high incidence of infection seen in health care workers.

We sought to determine which standard decontamination techniques used in hospitals might be suitable for the task of sterilizing a variety of N95 masks without compromising their structural or functional integrity. We also sought to ensure that each technique was effective in eliminating any viable virus by potential penetration through the surface as might be seen with large contaminated droplet deposition. Efforts at ultraviolet light decontamination, for example, might only sterilize the mask surface without having an impact on virus particles that have been deposited deeper [4].

Our tests of decontamination effectiveness demonstrate that all decontamination methods assessed were highly effective in sterilizing all the N95 models. No viable virus (SARS-CoV-2 where logistically possible and VSV as a surrogate if necessary) was found on any experimentally contaminated mask following any decontamination procedure (autoclave, PAF, VHP, EtO gas, or LT-HPGP). This is an expected result but is useful in that previous studies have made the assumption that such techniques would necessarily be effective on N95 masks [4-7].

*Vesicular stomatitis virus*, a bullet shaped enveloped, negative-sense RNA virus of the *Rhabdoviridae* family that commonly infects animals [8], was used as a surrogate for SARS-CoV-2 for decontamination procedures (LT-HPGP and EtO) only available at our hospital. We could not validate SARS-CoV-2 against these two technologies because it is a Risk Group 3 virus, which cannot be manipulated outside CL3.

More importantly, our results clearly show that the use of individual N95 masks can potentially be extended several-fold without degradation of functional integrity. VHP [9] and PAF appear to be most effective across all masks. There is recent preprint data that supports the possibility using a slightly different technology named Hydrogen Peroxide Vapor (HPV) [5]. We demonstrated that the VHP method allows at least 10 cycles of decontamination without impairment of mask function. The disadvantage of VHP is its cost and limited availability.

Peracetic acid dry fogging is an attractive, mobile and affordable decontamination technology [10]. This validation study demonstrated its ability to decontaminate all tested masks successfully without affecting their functional efficiency up to 10 cycles (maximum cycles tested). Handling and storage of extremely corrosive liquid peracetic acid and the routine cleaning requirement of the nozzles immediately after fogging to prevent clogging are the two disadvantages of this system.

Low temperature hydrogen peroxide gas plasma is commonly used in most hospitals for decontamination of high value reusable equipment such as endoscopes [11]. However, we were able to demonstrate that the N95 masks tolerated one standard (47 min) cycle of treatment. With 2 or more cycles, quantitative fit testing was impaired for all but one of six masks assessed. We postulate that the high concentration of liquid hydrogen peroxide (approximately 60%) and its strongly charged ionized vapor state of this device may have neutralized the filter media’s electrostatic charge, which is critical in trapping airborne particulates.

Ethylene oxide gas treatment is an older method of decontaminating materials [12]. The process is somewhat more complex than others and there can be safety concerns in that the gas is flammable, explosive and potentially carcinogenic [13]. A prolonged period of aeration following item exposure to the gas is required to eliminate chemical residue. This results in a considerably long cycle time of more than 20 hours compared to an hour or less for other decontamination methods. Despite these drawbacks, some institutions in poorly-resourced settings may not have LT-HPGP or VHP. For that reason, our finding that all four mask models tolerate at least 3 cycles of EtO decontamination without significant structural or functional deterioration may be useful. However, we would recommend against the use of this approach unless and until there is advanced testing to ensure that all traces of ethylene oxide and its related byproducts are entirely eliminated with sufficient aeration [14].

Finally, as expected, standard autoclaving is effective in eliminating any viable virus. Surprisingly, however, 4 of the 6 assessed respirator mask models tolerated up to 10 cycles while maintaining structural and functional integrity according to our testing. Although all masks maintained integrity after one autoclave cycle, the more rigid, molded 3M 1860 and 8210 models demonstrated loss of function with more than a single autoclave cycle. The other layered fabric, pleated models all retained integrity with up to 10 autoclave cycles (maximum number of cycles tested). This finding could be highly relevant to institutions in poorly-resourced areas of the world in that autoclaves would be expected to be available in any established hospital or major medical clinic around the world. Unfortunately, we were unable to examine the differences in mask materials and construction that might contribute to the failure of the 3M 1860 and 8210s model compared to the others due to the proprietary nature of the technology. It is worthwhile investigating autoclave process further using reduced temp and/or time conditions to see if 1860 and 8210 model masks can be decontaminated without losing their integrity upon repeat cycles.

Single use of N95 masks for each patient encounter is ideal and recommended; unfortunately, the resource stress due to the current COVID-19 crisis has breached this ideal. According to public reporting, extended use and re-use of N95 masks has become common in hospitals in areas where SARS-CoV-2 is high. This risks functional failure of N95 masks, spread of infection to wearers and increased risk of virus transmission from health care workers to others. Our data suggests that all decontamination methods are effective for at least one decontamination cycle without loss of structural integrity. However, neither LT-HPGP nor EtO gas are recommended at this time due to limited tolerance of N95 masks tested to repeat cycles, prolonged cycle times and/or potential toxicity. Peracetic acid dry fogging, VHP and autoclaving can be used to decontaminate N95 masks through multiple cycles without loss of function. Although VHP has limited availability and increased cost to acquire, dry fogging system can be easily procured and operated in comparison. Autoclaves can be used on a subset of N95 mask types and may be easily accessed by any healthcare institution when N95 mask shortages occur.

Although we tested the functionality of decontaminated masks via quantitative fit testing, our testing cannot take into account the respirator’s ability to withstand the rough handling that extended wear by health care workers, with stress and perspiration can inflict. Another limitation of this study is that our findings may or may not apply to other types of N95 masks. We also could not distinguish whether failure of fit or failure of filtration efficiency lead to the failings of those masks upon treatment by LT-HPGP or autoclave treatments.

Nonetheless, it is reassuring that the practice of appropriate decontamination and subsequent re-use of N95 mask should not pose a health risk to the already taxed health care workers.

## Data Availability

all manuscript data is available for review.

